# Service delivery in acute ischemic stroke patients: Does sex matter?

**DOI:** 10.1101/2023.06.19.23291634

**Authors:** Friedrich Medlin, Davide Strambo, Dimitris Lambrou, Valeria Caso, Patrik Michel

## Abstract

**Background:** Women with acute ischemic stroke (AIS) are older and have higher preexisting handicap than men. Given that these factors do not fully explain their poorer long-term outcomes, we sought to investigate potential sex differences in the delivery of acute stroke care in a large cohort of consecutive AIS patients.

**Methodss:** We analyzed all patients from the Acute STroke Registry and Analysis of Lausanne (ASTRAL) from 03/2003-12/2019. Multivariate analyses were performed on acute time metrics, revascularization therapies, ancillary exams for stroke work-up, subacute symptomatic carotid artery revascularization, frequency of change in goals of care (palliative care) and length of hospital stay.

**Results:** Of the 5347 analyzed patients, 45% were biologically female and the median age was 74.6 years. After multiple adjustments, female sex was significantly associated with higher onset-to-door (adjusted hazard ratio (aHR):1.09, 95% confidence interval (CI) 1.04-1.14) and door-to-endovascular-puncture intervals (aHR:1.15, 95%CI:1.05-1.25). Women underwent less diagnostic exams (adjusted odds ratio (aOR): 0.94, 95%CI:0.85-1.04), fewer subacute carotid revascularizations (aOR:0.69, 95% CI:0.33-1.18) and had longer hospital stays (aHR:1.03, 95%CI:0.99-1.07), but these differences were not statistically significant. We found no differences in the rates of acute revascularization treatments, or in the frequency of change of goals of treatments.

**Conclusions:** In this retrospective analysis of a large, consecutive AIS cohort, female sex was associated with unfavorable pre- and inhospital time metrics, and lesser diagnostic exam and carotid revascularization rates. Such indicators of less effective stroke care delivery may contribute to the poorer long-term functional outcomes in female patients and require further attention.

## Introduction

While there is mounting evidence that women are older and have higher preexisting handicap and comorbidities at stroke onset than men ^1–3^, this only partially explains the poorer long-term functional outcome in female stroke patients. Possibly, acute stroke care treatment, work-up, and secondary prevention could differ between sexes and contribute to these findings and more data are needed on this latter aspect of stroke management as well as on care delivery in the acute and subacute phases. Considering the expected age-dependent increase in stroke incidence in women in the next decades, this issue becomes even more important. In this quality assurance project, we sought to investigate potential sex differences in care delivery in acute ischemic stroke (AIS) in a large consecutive cohort of patients treated at our institution.

## Methods

### Patients and data collection

We reviewed retrospective data from March 2003 to December 2019 of all patients in the prospectively constructed Acute STroke Registry and Analysis of Lausanne (ASTRAL).^4^ ASTRAL collects data from all AIS patients admitted to the stroke unit and/or intensive care unit of Lausanne University Hospital (CHUV) within 24 hours of last known well time. Our center’s patient population consists of one-third distant referrals from community hospitals and stroke units mainly addressed for acute revascularization treatment, and about two-thirds of patients from the primary catchment area.

We recorded demographic data, vascular risk factors (arterial hypertension, smoking, atrial fibrillation, valve replacements, dyslipidemia, coronary artery disease and diabetes mellitus), multiple other comorbidities (like a history of renal disease, cancer, dementia, and depression) and previous cerebrovascular events. We collected medications at the time of stroke and the pre-stroke modified Rankin score (mRS). The National Institutes of Health Stroke Score (NIHSS) was performed or supervised on arrival by NIHSS-certified personnel.

Neuroimaging was CT-based up to 4/2018, after which MRI became the initial imaging modality of choice. CT-or MR-arteriography and perfusion imaging for supratentorial strokes were performed whenever possible.

Stroke mechanism was classified according to TOAST ^5^. The rest of the acute stroke management and secondary prevention followed European Stroke Organisation guidelines in use at the time of stroke. Rankin-certified personnel assessed mRS at 7 days, and then at 3 months in the outpatient clinic (or by a structured telephone interview for patients unable to attend the 3-month outpatient clinic).

### Outcomes

We chose eight outcomes to assess potential differences in care delivery between female and male stroke patients:

- Onset-to-door (OTD) time analyzed as a continuous variable, i.e., the time between the reported symptom onset (or last proof of good health in unknown and sleep-onset patients) and arrival at our tertiary hospital
- Proportion of acute revascularization treatments (intravenous thrombolysis (IVT) and/or endovascular treatment (EVT) in the first 24 hours. The decision to perform acute revascularization treatment followed the written multidisciplinary hospital guidelines that mirror Swiss ^6^ and European recommendations. ^7^
- Times to acute revascularization treatment, analyzed as continuous variables:
  ∘ Door-to-needle time in IVT, and/or
  ∘ Door-to-puncture (DTP) time in EVT
- Diagnostic exam score: we attributed one point for each of the following eight ancillary diagnostic exams performed during the subacute hospital phase in our stroke center: neurovascular ultrasound (cervical arteries with transcranial Doppler whenever a bone window was present), subacute cranial CT, subacute cranial MRI, subacute cerebral CTA or MRA (with or without cervical arteries), transthoracic and/or transesophageal echocardiography (TTE, TEE), and patent foramen ovale (PFO) detection by TTE and/or TEE with microbubble injection. The decision to perform these exams was taken by the treating physician in the stroke unit.
- Carotid revascularization treatments (CEA and/or CAS) within one month for a symptomatic stenosis ≥ 50%. The decision to perform these treatments was based on our published multidisciplinary hospital guidelines that are subject to Swiss ^8^ and European ^9^ recommendations at the time of the intervention. In summary, the preferred intervention was CEA; CAS was offered to patients with anatomical situations unsuitable for CEA, restenosis after previous revascularizations, post-radiation stenosis, and on demand from patients aged below 70 years.
- Change in goals of care (CGC) towards a palliative care attitude during the hospitalization for the index stroke. Patients or their legal representatives and the neurologist in charge addressed a CGC based on clinical variables such as initial course, comorbid conditions, pre-stroke disability, prior patient directives, or presumed wishes of the patient. Any CGC decision was registered in the patient’s medical record and in ASTRAL, including the time from stroke onset.
- Length of hospital stay, analyzed as a continuous variable. The onset of the hospital stay was defined as the time of arrival at the hospital of patients admitted thereafter to the stroke unit and/or intensive care unit of our institution (for in-hospital stroke, it was defined as the last proof of good health). The end of the hospital stay coincided with either discharge from the stroke unit and/or intensive care unit to another institution, or to another in-house acute medical service.

We applied the STROBE method (Strengthening the Reporting of Observational Studies in Epidemiology).^10^

## Ethical considerations

ASTRAL is registered in our institution as a clinical and research databank and follows the institutional regulations on clinical and research registries. Patients received a written document from the hospital that their routinely collected clinical data may be used for quality and scientific purposes, and we did not consider a patient’s decision to opt out from data analysis as this was a quality assurance project of the treatment practice in our institution. This type of analysis that aims to evaluate treatment efficiency and safety for quality purposes falls outside of the Swiss Human Research Act of 2011.

## Data availability

The raw, anonymized data that support the findings of this study are available from the corresponding author upon reasonable request and after signing a data transfer and use agreement (DTUA). If such data are then used for a publication, the publication methods should be communicated, and internationally recognized authorship rules should be applied.

## Statistical Analyses

Study participants’ data were analyzed according to sex classification, as shown in figure 1. We assessed demographic, clinical, and radiological variables from the acute phase of stroke (<24 hours) recorded in the ASTRAL database. Continuous variables were summarized as median with interquartile range (IQR) and categorical variables as percentages and numbers.

Initially, univariate comparisons were performed according to sex and the unadjusted odds ratios (ORs) were evaluated. Subsequently, multiple regression analyses of the aforementioned eight main outcomes were conducted to identify independent associations with sex, after adjusting for the following 23 covariates: age, ethnicity (Caucasian vs. all others), prehospital referral pattern (patients coming from primary vs. secondary hospital catchment areas), time of symptom onset to hospital admission, history of prior cerebrovascular events, stroke severity, pre-hospital admission mRS, cerebrovascular risk factors (hypertension, diabetes, hyperlipidemia, smoking, atrial fibrillation, alcohol abuse, body-mass index), peripheral artery disease, coronary artery disease, cardiac heart valves, cardiac ejection fraction, migraine, active cancer, depression and psychosis. Additional adjustments were made for specific analyses: OTD time to assess the proportion of acute revascularization treatment (IVT and/or EVT), OTD time and carotid stenosis grade to evaluate carotid revascularization treatments, and OTD time to assess length of hospital stay and a CGC.

For *continuous* response variables, a Cox proportional hazards model was employed to associate covariates with the response. This approach has the advantage that no distributional assumptions are made for the continuous response. For *categorical* outcomes, with limited values, a proportional odds model was used to identify important links between response and background information.

Missing data were imputed using multivariate imputation by chained equations (MICE) methodology. Five imputed datasets were generated. Stepwise selection methods were employed to identify significant associations of the selected covariates with the specific outcome on each imputed dataset. Results of the five imputed analyses were combined appropriately to formulate the final analysis results that were given as adjusted odds ratios (aORs) with confidence intervals (CIs) for categorical and ordinal outcomes, and adjusted hazard ratios (aHR) with CIs for continous outcomes. All statistical analyses were conducted using the R statistical software version 4.4.2 (R Core Team 2017).

## Results

Retrospective data from 5347 patients who fulfilled the inclusion criteria were analyzed (figure 1). Of these patients, 45% were female, 3.8% were of non-Caucasian ethnicity and the median age was 74.6 years. The proportion of women in the population referred from the secondary catchment area was 30.4%, lower than the proportion of men (34.7%). Detailed sex-specific description of the cohort is described in table 1 and supplemental table 1; it resembles closely our previous sex-specific analysis concentrating on clinical presentations and long-term outcomes ^2^.

**Table 1:** Overall and sex-specific unadjusted baseline characteristics of the population. Values are expressed as medians and interquartile range for continuous variables, and absolute counts and percentages for categorical variables unless otherwise stated.

| Variable | Overall<br>N=5.347 | Female sex<br>N=2.395 | Male sex<br>N=2.952 |
| --- | --- | --- | --- |
| *Demographics and major vascular risk factors* |  |  |  |
| Age | 74.6 (19.7) | 78.1 (17.0) | 71.2 (19.9) |
| Hypertension | 3944 (73.8) | 1802 (75.3) | 2142 (72.6) |
| Diabetes | 1057 (19.8) | 387 (16.2) | 670 (22.7) |
| Hyperlipidemia | 4068 (76.2) | 1768 (73.9) | 2300 (78.0) |
| Smoking (current or stopped < 2 years) | 1219 (23) | 418 (17.6) | 801 (27.4) |
| Atrial fibrillation | 1621 (30.3) | 846 (35.4) | 775 (26.3) |
| *Prehospital and admission data* |  |  |  |
| Known time of stroke onset | 3623 (67.8) | 1543 (64.5) | 2080 (70.5) |
| Stroke present at wake-up | 1313 (24.6) | 602 (25.1) | 711 (24.1) |
| mRS pre-stroke | 1.0 (2.0) | 1.0 (2.0) | 0.0 (1.0) |
| Previous ischemic cerebral event | 951 (17.8) | 387 (16.2) | 564 (19.1) |
| OTD time (hours) | 3.1 (8.7) | 3.4 (9.9) | 3.0 (7.7) |
| Patient referred from outside the primary catchment area | 1751 (32.8) | 727 (30.4) | 1024 (34.7) |
| NIHSS on admission | 6.0 (11.0) | 7.0 (12.0) | 6.0 (10.0) |
| *Acute treatment* |  |  |  |
| IVT (with or without EVT) | 1268 (23.7) | 543 (22.7) | 725 (24.6) |
| EVT (with or without preceding IVT) | 812 (15.2) | 365 (15.2) | 447 (15.1) |
| Door-to-IVT time (hours) | 0.7 (0.7) | 0.7 (0.7) | 0.7 (0.7) |
| Door-to-puncture time (hours) | 1.6 (1.1) | 1.6 (1.0) | 1.5 (1.1) |
| Symptomatic extracranial carotid stenosis 50-99% | 795 (17.4) | 279 (13.9) | 516 (20.2) |
| *Other outcome measures* |  |  |  |
| Subacute carotid revascularization procedure at 12 months | 168 (3.2) | 39 (1.7) | 129 (4.4) |
| Number of subacute diagnostic exams (diagnostic exam score) | 3.0 (2.0) | 3.0 (2.0) | 3.0 (2.0) |
| Length of hospital stay (days) | 8.0 (8.0) | 9.0 (8.0) | 8.0 (8.0) |
| Change in goals of care during acute hospitalization | 561 (10.5) | 294 (12.3) | 267 (9.1) |

mRS, modified Rankin Scale; NIHSS, National Institutes of Health Stroke Scale; IVT, Intravenous thrombolysis; EVT, endovascular treatment.

Unadjusted comparisons (see table 1 and supplemental table 1) showed a significantly higher OTD time (3.4 vs. 3.0 hours; HR 1.02; CI95% 1.01-1.03), a lower proportion of IVT (22.7% vs. 24.6%; OR 0.9; CI95% 0.79-1.02) and a slightly higher proportion of EVT treatments (15.2% vs. 15.1%; OR 0.98; CI95% 0.84-1.14) for women compared to men. Furthermore, women had fewer diagnostic exams (median number 3 vs. 3, OR 0.9 CI95% 0.87-0.93), fewer subacute carotid revascularization treatments (1.7% vs. 4.4%; OR 0.36 CI95% 0.25 - 0.52), longer lengths of hospital stay (9 vs. 8 days; HR 1.01 CI95% 1.00 - 1.01) and a higher proportion of change in goals of care (12.3% vs. 9.1%; OR 1.40 CI95% 1.18 - 1.67) compared to men.

In the adjusted multivariate analysis (see table 2) female sex remained significantly associated with higher OTD (aHR: 1.09, 95%CI 1.04-1.14; p<0.01) and DTP times (aHR: 1.15, 95%CI 1.05-1.25; p<0.01). While not statistically significant, we observed a lower number of diagnostic exams (aOR: 0.94, 95%CI 0.85-1.04; p=0.21), fewer subacute carotid revascularization treatments (aOR: 0.69 95%CI 0.33-1.18; p=0.147) and longer hospital durations (aHR: 1.03, 95%CI 0.99-1.07; p=0.19) in women. Further, in the multivariate analysis we no longer saw trends in the proportions of IVT and/or EVT or changes in goals of treatments between male and female sex patients.

**Table 2:** Summary of the multivariate analysis results for each of the eight sex-specific outcomes (reference male sex)

| Outcome | Unadjusted ratio* | 95% CI | P value unadjusted | Adjusted ratio* | 95% CI | P value adjusted |
| --- | --- | --- | --- | --- | --- | --- |
| Onset-to-door time (hours) | 1.02 | 1.01 - 1.03 | <0.01 | 1.09 | 1.04-1.14 | <0.01 |
| Diagnostic exam score (0-8) | 0.91 | 0.88 - 0.94 | <0.01 | 0.85 | 0.85-1.04 | 0.21 |
| Acute revascularization treatment, i.e., IVT and/or EVT (%) | 1.09 | 0.91 - 1.3 | 0.26 | 1.05 | 0.91-1.22 | 0.48 |
| Door-to-needle time for IVT (hours) | 0.99 | 0.97 - 1.01 | 0.28 | 0.95 | 0.86 -1.05 | 0.34 |
| Door-to-puncture time for EVT (hours) | 1.01 | 0.95 - 1.07 | 0.75 | 1.15 | 1.05 -1.25 | <0.01 |
| Subacute revascularization for symptomatic carotid stenosis (50-99%) | 0.36 | 0.25 - 0.52 | <0.01 | 0.63 | 0.33-1.18 | 0.15 |
| Length of hospital stay (days) | 1.01 | 1.00 - 1.01 | 0.6 | 1.03 | 0.99-1.07 | 0.19 |
| Change in goals of care | 1.4 | 1.18 - 1.67 | <0.01 | 0.99 | 0.8-1.23 | 0.92 |

\* Odds ratio for categorical and ordinal outcomes, and hazard ratio for continuous outcomes
CI, confidence interval; IVT, intravenous thrombolysis; EVT, endovascular treatment. For detailed results of each outcome, see supplemental tables. For factors used for adjustment, see methods section.

## Discussion

In this retrospective study of 5347 consecutive AIS patients admitted to a comprehensive stroke center, we found that female sex was significantly associated with longer OTD and DTP times. Additionally, we noted numerically fewer diagnostic exams in women during the acute hospital stay, fewer subacute carotid revascularization treatments and longer hospital durations, but these differences did not reach statistical significance. Altogether, these indicators of less effective stroke care delivery may contribute to the poorer long-term functional outcomes in female patients ^2, 11^ and warrants further attention.

Existing literature focusing on sex differences in acute stroke care and service delivery is limited and yields inconsistent results. An older analysis from the GWTG-Stroke program ^12^ and a recent large retrospective study in China ^1^ did not show significant sex differences in acute management and intrahospital and discharge performance measures. A pooled analysis of five randomized controlled trials found that women were more likely to be admitted to an acute stroke unit ^14^, whereas other studies from France and Canada showed the contrary.^14, 15^

This same pooled analysis showed that women were less likely to benefit from intensive care treatment and acute interventions such as intubation ^13^, nasogastric feeding ^13^, and treatment for fever ^13^. The latter finding was confirmed by a recent nationwide study from Canada demonstrating that women and older patients (≥80 years) had lower odds of receiving life-sustaining care, but disparities in intensive care unit admission narrowed over time.^16^

Regarding prehospital timing, several other studies reported longer onset-to-hospital times for women ^17–22^. whereas others did not find a sex-related difference. ^23, 24^ One plausible reason for increased pre-hospital times is that women are more often widowed and living alone, ^25^ leading to a later activation of emergency medical services. ^21^ Another explanation might be more non-focal, atypical, and non-traditional symptoms in acute stroke presentations in women, leading to under-recognition of stroke and non-activation of stroke alert pathways. ^26, 27^

Regarding intravenous thrombolysis, our results confirmed those of other studies showing comparable treatment rates and time metrics for both sexes. ^1, 22, 23, 28, 31^ However, one study of Danish public registries ^21^ and a recent meta-analysis of 17 studies showed 13% lower unadjusted odds of receiving IV rtPA in women. ^30^ Reassuringly, such differences seem to decrease towards more sex equity over time: a recent nationwide analysis in the US showed that in the last decade, utilization of rtPA increased at a faster pace in women compared to men. ^31^ This trend was confirmed by epidemiological data from France ^15^ and by the aforementioned meta-analysis by Strong and co-workers. ^30^ Recent data from Catalonia for the period from 2016 to 2019 showed even a higher proportion of acute reperfusion therapies in women.^24^

Regarding the rate of EVT, we found no differences between sexes after adjusting for several factors, whereas other studies showed higher EVT rates in women.^14, 30, 32^ Given that several of these other studies ^14, 32^ adjusted their comparisons with multiple co-variates, there is no simple explanation for these findings such as higher admission NIHSS scores or rates of atrial fibrillation.^2^

We found longer door-to-groin times in women treated with endovascular therapies, despite adjusting for prehospital delays. This contrasts with other studies that found similar intrahospital times for EVT. ^14, 24, 33, 34^ Our finding does not have an obvious explanation, but a possible explanation is a cognitive sex bias.

Regarding the in-hospital stroke work-up in the subacute phase, our results showed that we performed numerically fewer diagnostic exams in women, but this did not reach statistical significance. Again, prior literature is divided when reporting on sex differences in diagnostic evaluation of stroke, with evidence for less extracranial magnetic resonance angiography (MRA),^23^ echocardiography and carotid evaluation in women ^35, 36^ while, on the contrary, older populations ^37^ and multicentric studies in the US and Canada did not demonstrate significant sex inequities for these exams. ^38, 39^

A further main result of our study was that after adjusting for age and stenosis grade, we observed numerically fewer subacute carotid vascularization procedures in women, not reaching statistical significance. This result may be explained by evolving sex-specific treatment recommendations for symptomatic carotid stenosis. ^40, 41^ Some studies show a higher stroke risk in male carotid stenosis patients compared to women, probably due to higher plaque instability ^42^, but endarterectomy risk seems similar ^43^, and the meta-analysis of randomized trial data does not show a differential effect of carotid revascularization depending on biological sex. ^41^

As for the lengths of hospital stay, the longer hospital duration for women was not significant after adjusting for the known higher stroke severity and pre-stroke handicap.^2^ Longer hospitalization in women has been reported by Somerford et al ^44^, possibly related to the more frequent absence of a spouse at home who could participate in the care.

Finally, regarding the complex question of sex disparities in CGC in AIS, we did not observe a significant difference in CGC decisions in our adjusted analysis. This confirms what we found in our recent analysis that identified several powerful predictors of GCS, but not sex.^45^

Limitations of our work are its retrospective, non-randomized design in a tertiary stroke center with approximately one-third of patients being referred from other hospitals or stroke units. As a quality assurance project in a single institution, it may not be applicable to other populations, in particular, non-Caucasian and younger populations. As stated above, we did not measure several variables that may have confounded our main outcomes, such as educational level, extent of social networks, isolation and loneliness, and socioeconomic and sociocultural status.

The strengths of our work are the consecutive nature of the collected data over a long period with pre-specified and standardized data collection, and rigorous adjustment of multiple confounders for the eight main outcome analyses.

In conclusion, this study suggests that female patients with AIS arriving at a comprehensive stroke center have longer OTD and DTP times while other indicators for care delivery are not statistically different. Some of these observations may contribute to the explanation of poorer long-term functional outcomes in female patients and require further attention.

## Acknowledgements

We thank Mrs. Price Hirt, PhD, for critical review and English-language correction of the manuscript.

## Author contributions

Friedrich Medlin: Study concept and design, data analysis and interpretation, drafting of the manuscript.

Dimitris Lambrou: Statistical analysis, software support, drafting of the manuscript. Davide Strambo: Data collection, data analysis and interpretation.

Valeria Caso: Study concept and design, critical revision of the manuscript.

Patrik Michel: Study concept and design, data collection, collection and handling of funding, critical revision of the manuscript, supervision of study.

## Non-standard Abbreviations and Acronyms

AIS: Acute ischemic Stroke
CGC: Change in goals of care
CTA: Computed tomography angiography
DTP: Door-to-puncture
EVT: Endovascular treatment
IVT: Intravenous thrombolysis
NIHSS: National Institutes of Health Stroke Score
MRA: Magnetic Resonance Angiography
mRS: Modified Rankin score
OTD: Onset-to-door
PFO: Patent foramen ovale
TEE: Transesophageal echocardiography
TTE: Transthoracic echocardiography

## Supplemental material

**Supplemental figure 1.**
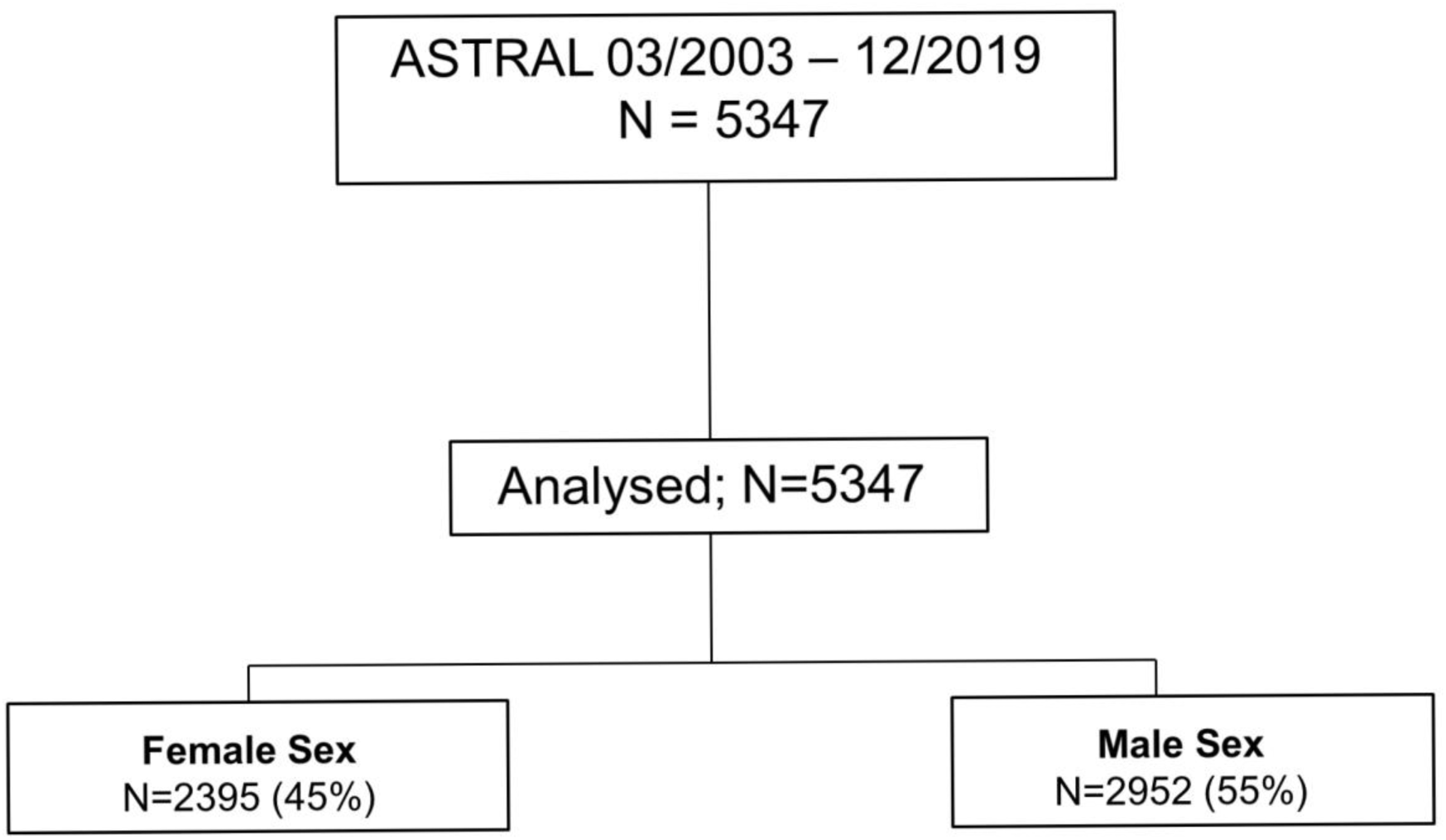
Study flow diagram

**Supplemental table 1:** Additional overall and sex-specific unadjusted baseline characteristics of the population. Values are expressed as medians and interquartile range for continuous variables, and absolute counts and percentages for categorical variables unless otherwise stated.

| Variable | Overall<br>N=5.347 | Female sex<br>N=2.395 | Male sex<br>N=2.952 |
| --- | --- | --- | --- |
| *Demographics, cardiovascular risk factors and comorbidities* |  |  |  |
| Ethnicity (Caucasian) | 5129 (96.2) | 2301 (96.2) | 2828 (96.1) |
| Pre-stroke mRS 3-5 | 685 (12.8) | 397 (16.6) | 288 ( 9.8) |
| Coronary artery disease | 1041 (19.5) | 337 (14.1) | 704 (23.9) |
| Heart valves | 200 (3.7) | 81 (3.4) | 119 (4.0) |
| Ejection fraction < 35% | 294 (5.5) | 91 (3.8) | 203 (6.9) |
| Symptomatic peripheral artery disease | 381 (7.2) | 133 (5.6) | 248 (8.4) |
| Active cancer | 329 (6.2) | 124 (5.2) | 205 (7.0) |
| Migraine with or without aura | 284 (5.4) | 183 (7.7) | 101 (3.5) |
| Alcohol abuse (current) | 605 (11.4) | 148 (6.2) | 457 (15.6) |
| Body-mass index (kg/m<sup>2</sup>) | 25.0 (5.0) | 24.0 (7.0) | 26.0 (4.0) |
| Depression | 229 (4.3) | 126 (5.3) | 103 (3.5) |
| Psychosis | 499 (9.4) | 291 (12.2) | 208 (7.1) |
| *Medications prior to stroke onset* |  |  |  |
| Antiplatelets | 2028 (37.9) | 834 (34.8) | 1194 (40.4) |
| Anticoagulants | 727 (13.7) | 345 (14.5) | 382 (13.0) |
| Antihypertensive treatment | 3255 (61) | 1539 (64.4) | 1716 (58.2) |
| Lipid-lowering drugs | 1628 (30.5) | 620 (25.9) | 1008 (34.2) |
| Antidiabetic treatment | 754 (14.1) | 266 (11.1) | 488 (16.6) |
| *Initial acute imaging modality* |  |  |  |
| CT | 4524 (84.6) | 2032 (84.9) | 2492 (84.4) |
| MRI | 662 (12.4) | 295 (12.3) | 367 (12.4) |
| None within 24h of stroke onset | 159 (3.0) | 67 (2.8) | 92 (3.1) |
| Door-to-imaging time (hours) | 1.0 (3.4) | 1.2 (3.5) | 0.8 (3.2) |
| Onset-to-imaging time (hours) | 4.4 ( 9.8) | 4.9 (10.6) | 4.0 ( 8.7) |
| *Initial acute vascular imaging modality* |  |  |  |
| None | 587 (11.0) | 307 (12.8) | 280 ( 9.5) |
| CTA | 4102 (76.8) | 1800 (75.3) | 2302 (78) |
| MRA | 654 (12.2) | 285 (11.9) | 369 (12.5) |
| *Acute CT/MRI perfusion imaging* |  |  |  |
| No good-quality perfusion study in acute phase<br>(not done/technical failure/other) | 1385 (27.7) | 631 (28.4) | 754 (27.1) |
| Good quality perfusion study available | 3644 (72.5) | 1607 (72) | 2037 (73.0) |
| Renal exclusion criteria | 223 (4.4) | 116 (5.2) | 107 (3.8) |
| Contrast allergy | 55 (1.1) | 32 (1.4) | 23 (0.8) |
| Technical reasons (patient or radiological) | 229 (4.6) | 84 (3.8) | 145 (5.2) |
| Other reasons | 487 (9.7) | 203 (9.1) | 284 (10.2) |
| *Stroke mechanism* |  |  |  |
| Atherosclerotic | 783 (14.7) | 242 (10.2) | 541 (18.5) |
| Cardioembolic | 1766 (33.2) | (37.3) | 879 (30) |
| Microangiopathic | 573 (10.8) | 249 (10.5) | 324 (11.1) |
| Unknown | 490 (9.2) | 201 (8.4) | 289 (9.9) |
| Multiple or co-existing causes | 1358 (25.6) | 669 (28.1) | 689 (23.5) |
| *Stroke localization* |  |  |  |
| Anterior circulation stroke | 3610 (68.8) | 1686 (71.5) | 1924 (66.6) |
| Posterior circulation stroke | 1288 (24.6) | 507 (21.5) | 781 (27.1) |
| Anterior and posterior circulation strokes | 131 (2.5) | 64 (2.7) | 67 (2.3) |
| Undetermined localization | 216 (4.1) | 101 (4.3) | 115 (4.0) |
| Right brain | 2159 (40.6) | 995 (41.7) | 1164 (39.6) |
| Left brain | 2593 (48.7) | 1168 (49) | 1425 (48.5) |
| Bilateral | 525 (9.9) | 201 (8.4) | 324 (11) |
| Undetermined side | 47 (0.9) | 22 (0.9) | 25 (0.9) |
| *Acute treatment* |  |  |  |
| Onset-to-IVT time (hours) | 2.3 (1.5) | 2.3 (1.6) | 2.2 (1.6) |
| Onset-to-puncture time (hours) | 4.0 (3.3) | 4.4 (4.0) | 3.9 (3.0) |
| Groin-puncture-to-recanalization time (hours) | 0.8 (0.8) | 0.8 (1.0) | 0.8 (0.8) |
| Door-to-recanalization time (hours) | 2.4 (1.6) | 2.6 (1.6) | 2.4 (1.6) |
| Onset-to-recanalization time (hours) | 5.2 (3.4) | 5.4 (4.3) | 4.9 (3.2) |
| *Other outcome measures* |  |  |  |
| Decompressive craniectomy for ischemic mass effect | 47 (0.9) | 15 (0.6) | 32 (1.1) |
| PFO closure in first 12 months | 74 (4.1) | 29 (3.7) | 45 (4.5) |
| Intensive care stay if applicable (days) | 487 (9.2) | 183 (7.7) | 304 (10.4) |
| mRS at 3 months (median) | 2.0 (3.0) | 2.0 (3.0) | 2.0 (2.0) |
| mRS 0-2 at 3 months | 2939 (58.9) | 1175 (52.7) | 1764 (63.9) |
| Mortality at 3 months | 769 (14.4) | 398 (16.6) | 371 (12.6) |
| Further cerebrovascular events over 3 months | 286 (6.6) | 124 (6.4) | 162 (6.8) |
| *Discharge destination* |  |  |  |
| Home | 1954 (36.8) | 770 (32.4) | 1184 (40.4) |
| Rehabilitation or other acute care hospital | 2884 (54.4) | 1374 (57.9) | 1510 (51.5) |
| In-hospital death | 468 (8.8) | 229 (9.7) | 239 (8.1) |

OR indicates odds ratio; CI, confidence interval; mRS, modified Rankin Scale; IVT, Intravenous thrombolysis; EVT, endovascular treatment; PFO, patent foramen ovale; CTA, Computed Tomography Angiography; MRA, Magnetic Resonance Angiography; TTE, Transthoracic Echocardiogram; TEE, Transesophageal Echocardiogram.

**Supplemental table 2:**
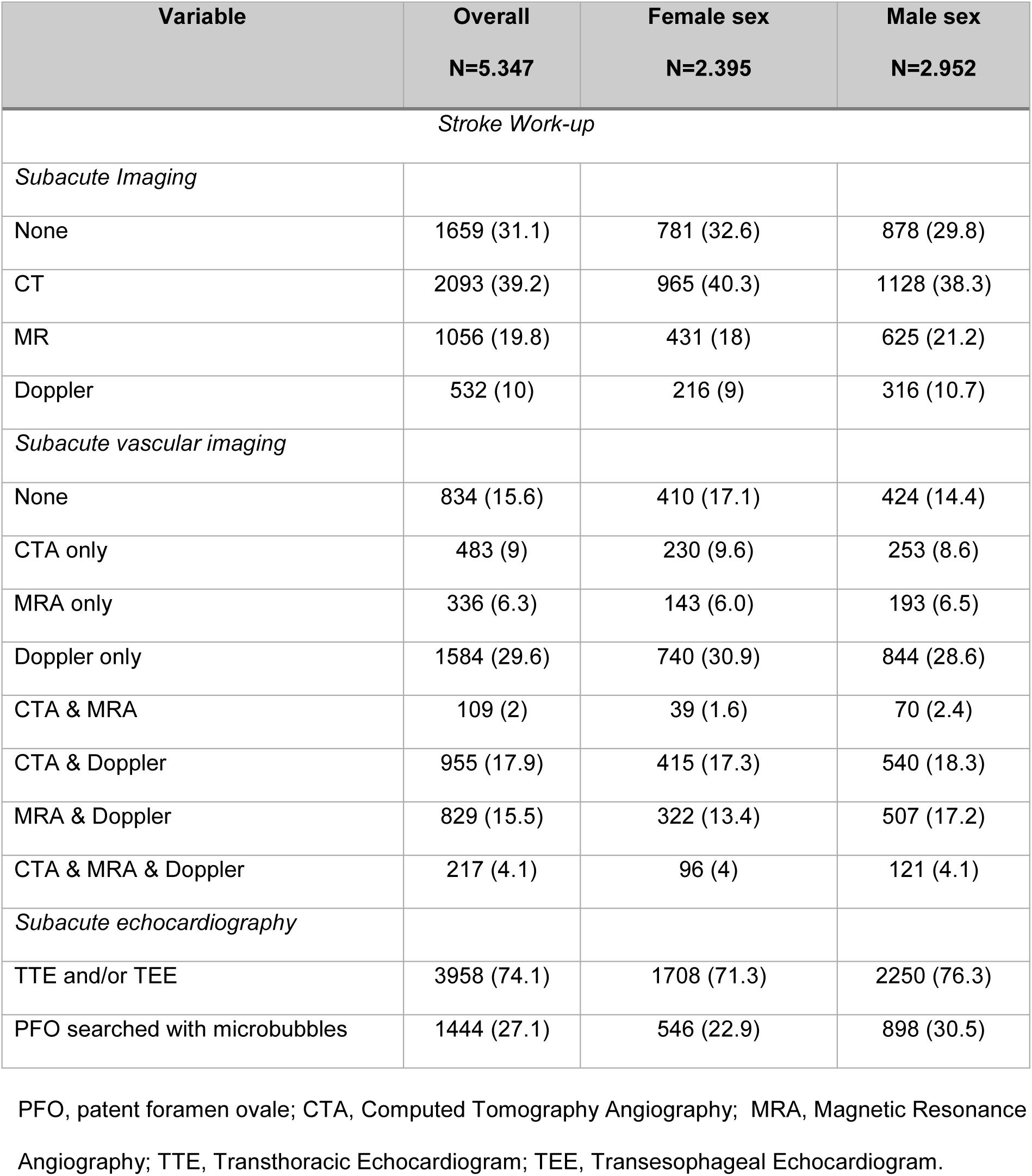
Overall and sex-specific unadjusted number of diagnostic exams in the subacute phase of the hospitalization. Values are expressed as absolute counts and percentages for categorical variables unless otherwise stated.

**Supplemental table 3:** Significant variables in the MVA of OTD time

| Variable | aHR | CI 95% | P value % |
| --- | --- | --- | --- |
| Sex (female) | 1.09 | 1.04 - 1.14 | 0.1 |
| Onset (known) | 0.29 | 0.27 - 0.30 | 0.0 |
| Stroke referral (outside) | 0.93 | 0.89 - 0.98 | 0.9 |
| Hypertension | 1.08 | 1.02 - 1.14 | 0.6 |
| Atrial fibrillation | 0.87 | 0.83 - 0.92 | 0.0 |
| Cardiac Valvulopathy | 0.85 | 0.74 - 0.97 | 1.6 |
| BMI | 0.99 | 0.99 - 1.00 | 0.6 |
| NIHSS admission | 0.97 | 0.97 – 0.98 | 0.0 |

aHR indicates adjusted hazard ratio; CI, confidence interval; BMI, Body Mass Index; NIHSS, National Institutes Health Stroke Scale. For other (non-significant) factors used for adjustment, see methods section.

**Supplemental table 4:** Significant variables in the MVA of diagnostic exam score

| Variable | aOR | CI 95% | P value % |
| --- | --- | --- | --- |
| Age | 0.96 | 0.96-0.97 | 0.0 |
| Onset (known) | 1.25 | 1.12 - 1.38 | 0.0 |
| Stroke referral (outside) | 1.61 | 1.45 - 1.78 | 0.0 |
| Cholesterol | 1.26 | 1.12- 1.41 | 0.0 |
| Symptomatic peripheral artery disease | 0.77 | 0.64 – 0.93 | 0.7 |
| Migraine | 1.58 | 1.27 - 1.96 | 0.0 |
| Previous ischemic cerebral event | 1.39 | 1.10 - 1.77 | 0.6 |
| Psychosis | 1.21 | 1.03 - 1.43 | 2.3 |
| Pre-hospital mRS | 0.87 | 0.83 - 0.91 | 0.0 |
| Sex (female) | 0.94 | 0.85 - 1.04 | 21.6 |

aOR indicates adjusted hazard ratio; CI, confidence interval; mRS, modified Rankin; NIHSS, National Institutes Health Stroke Scale.

**Supplemental table 5:** Significant variables in the MVA of acute revascularization treatment (IVT/EVT)

| Variable | aOR | CI 95% | P value % |
| --- | --- | --- | --- |
| Onset (known) | 1.22 | 1.02 - 1.47 | 3.4 |
| Stroke referral (outside) | 6.28 | 5.4 - 7.30 | 0.0 |
| Cholesterol | 1.68 | 1.42 - 1.99 | 0.0 |
| Cardiac Valvulopathy | 0.68 | 0.47 - 0.97 | 3.3 |
| Active Cancer | 0.70 | 0.52 - 0.93 | 1.5 |
| Migraine | 1.42 | 1.04 - 1.95 | 2.8 |
| Previous cerebrovascular event | 0.80 | 0.66 - 0.98 | 2.7 |
| Pre-hospital mRS | 0.85 | 0.79 - 0.90 | 0.0 |
| NIHSS admission | 1.10 | 1.09 - 1.11 | 0.0 |
| OTD time (h) | 0.81 | 0.79 - 0.82 | 0.0 |
| Sex (female) | 1.05 | 0.91 - 1.22 | 47.9 |

aOR indicates adjusted odds ratio; CI, confidence interval; mRS, modified Rankin; NIHSS, National Institute Health Stroke Scale. OTD; Onset-to-door time.

**Supplemental table 6:** Significant variables in the MVA of door-to-needle-time

| Variable | aHR | CI 95% | P value % |
| --- | --- | --- | --- |
| Age | 0.99 | 0.99 - 1.00 | 0.0 |
| Cardiac valvulopathy | 1.46 | 1.09 - 1.95 | 1.1 |
| OTD time | 1.23 | 1.2 - 1.27 | 0.0 |
| Sex (female) | 0.95 | 0.86 – 1.05 | 33.8 |

aHR indicates adjusted hazard ratio; CI, confidence interval; OTD; Onset-to-door time.

**Supplemental table 7:** Significant variables in the MVA of door-to-groin-time

| Variable | aHR | CI 95% | P value % |
| --- | --- | --- | --- |
| Sex (female) | 1.15 | 1.05 - 1.25 | 0.3 |
| Onset (known) | 0.66 | 0.58 - 0.76 | 0.0 |
| Stroke referral (outside) | 0.66 | 0.59 - 0.73 | 0.0 |
| Peripheral artery disease | 1.20 | 1.01 - 1.43 | 4.2 |
| Previous cerebrovascular event | 1.12 | 1.00 - 1.27 | 5.5 |
| OTD time (h) | 0.88 | 0.86 - 0.9 | 0.0 |

aHR indicates adjusted hazard ratio; CI, confidence interval; OTD; Onset-to-door time.

**Supplemental table 8:** Significant variables in the MVA of subacute revascularization at 6 months for symptomatic carotid stenosis (50-99%)

| Variable | aOR | CI 95% | P value % |
| --- | --- | --- | --- |
| Sex (female) | 0.48 | 0.28 - 0.81 | 0.6 |
| Cholesterol | 2.06 | 1.07 - 3.96 | 3.2 |
| Smoking | 1.81 | 1.15 - 2.85 | 1.1 |
| Coronary artery disease | 0.44 | 0.24 - 0.79 | 0.6 |
| Psychosis | 0.32 | 0.11 - 0.94 | 3.9 |
| NIHSS admission | 0.88 | 0.84 - 0.91 | 0.0 |

aOR indicates adjusted odds ratio; CI, confidence interval; NIHSS, National Institutes Health Stroke Scale.

**Supplemental table 9:** Significant variables in the MVA of length of hospital stay

| Variable | aHR | CI 95% | P value % |
| --- | --- | --- | --- |
| Sex (female) | 0.98 | 0.93-1.04 | 59 |
| Ethnicity (Caucasian) | 1.17 | 1.02-1.35 | 2.5 |
| Onset (known) | 1.09 | 1.03-1.15 | 0.4 |
| Stroke referral (outside) | 1.66 | 1.56-1.76 | 0.0 |
| Hypertension | 0.89 | 0.84-0.95 | 0.0 |
| Depression | 0.79 | 0.69-0.90 | 0.0 |
| Psychosis | 0.89 | 0.81-0.98 | 1.3 |
| Prehospital mRS | 0.92 | 0.9-0.95 | 0.0 |
| NIHSS admission | 0.98 | 0.97-0.98 | 0.0 |

aHR indicates adjusted hazard ratio; CI, confidence interval; NIHSS, National Institutes Health Stroke Scale.

**Supplemental table 10:** Significant variables in the MVA of change in goals of care decision

| Variable | aOR | CI 95% | P value % |
| --- | --- | --- | --- |
| Age | 1.04 | 1.03-1.05 | 0.0 |
| Sex (female) | 0.99 | 0.8-1.23 | 92.1 |
| IVT vs. none | 0.82 | 0.64-1.06 | 12.6 |
| EVT vs. none | 0.65 | 0.48-0.85 | 0.3 |
| Cholesterol | 0.65 | 0.52-0.82 | 0.0 |
| Coronary artery disease | 1.62 | 1.27-2.06 | 0.0 |
| Active cancer | 2.77 | 1.95-3.93 | 0.0 |
| Peripheral Artery Disease | 1.52 | 1.07-2.17 | 1.9 |
| Prehospital mRS | 1.36 | 1.25-1.49 | 0.0 |
| NIHSS admission | 1.18 | 1.16-1.2 | 0.0 |

aOR indicates adjusted odds ratio; CI, confidence interval; NIHSS, National Institutes Health Stroke Scale. IVT, Intravenous thrombolysis; EVT, endovascular treatment; mRS, modified Rankin.

